# Analgesic Efficacy and Perioperative Safety of the Erector Spinae Plane Block in Adult Cardiac Surgery

**DOI:** 10.1101/2025.08.28.25334223

**Authors:** Kanika Purohit, Timerjot Kaur, Abhishek Mehan, Rahul Chopra, Parag N Patel, Fnu Kalpana, Yashas Maragowdanahalli Somegowda, Ibrahim Siddiqui, Rakhshanda khan, Harshawardhan Dhanraj Ramteke, Ritesh Mate

## Abstract

**Introduction:** Severe post-sternotomy pain and the constraints of neuraxial techniques under anticoagulation highlight the need for opioid-sparing, feasible analgesia in adult cardiac surgery. The erector spinae plane (ESP) block—providing paravertebral spread with a favorable safety profile—may reduce pain and opioid consumption without compromising hemodynamic stability; here we assess its analgesic efficacy and perioperative safety.

**Methods:** A systematic search of various databases was conducted for Randomized Controlled Studies (RCTs) on erector spinae plane (ESP) block up to August 2025. Meta-analysis was performed using Stata 18.0, and ROBS 2.0 assessed risk of bias.

**Results:** A total of 2658 Studies were screened, out of which 24 RCTs were included in the meta-analysis. Total number of patients were 1687, out of which 1073 were male, 637 were female, average age was 55.72 ± 6.40 with average follow up hours were 44.75 ± 18.29. Total number of patients in treatment group were 856 and total number of patients in control group were 837. Resting VAS pain showed no difference at 0 h (MD −0.03, 95% CI −0.36 to 0.30) or 24 h (0.01, −0.32 to 0.34), a increase at 36 h (0.50, 0.08 to 0.92), and no difference at 72 h (0.27, −0.40 to 0.94). During coughing, VAS favored control at 0 h (0.76, 0.03 to 1.48), 24 h (1.12, 0.03 to 2.21), and 36 h (0.74, 0.06 to 1.43), but not 72 h (0.25, −0.82 to 1.32). Opioid use decreased at 36 h (−24.87, −34.27 to −15.48). Length of stay was unchanged (−0.21 days, −0.70 to 0.28). Adverse events did not differ. Risk of bias was low.

**Conclusion:** The ESP block is a safe, feasible, opioid-sparing adjunct with overall comparable pain scores and no increase in complications, supporting its use in adult cardiac surgery.

## 1. Introduction

Globally, more than one million patients undergo cardiac surgery each year, creating a large population exposed to severe post-sternotomy pain, prolonged intensive care monitoring, and extended hospitalization [1]. Effective perioperative analgesia is therefore not merely an adjunct to surgical care but a central determinant of recovery quality, time to extubation and mobilization, and overall resource utilization [2]. Although opioid-based regimens remain the most common pharmacologic strategy, their dose-dependent adverse effects— hemodynamic instability, ileus, nausea and vomiting, sedation, and respiratory depression—can paradoxically impede rehabilitation and delay discharge [3,4]. Of additional concern is the nontrivial incidence of new persistent opioid use following cardiac procedures, which elevates the importance of opioid-sparing pathways. Thoracic epidural analgesia, historically considered a reference technique for post-sternotomy analgesia, is constrained in contemporary cardiac practice by anticoagulation requirements, the risk—however rare—of epidural hematoma and other neuraxial complications, and practical barriers to routine placement in hemodynamically labile patients [5]. These limitations collectively underscore the need for analgesic strategies that deliver robust, reproducible pain control without compromising safety or feasibility in complex cardiothoracic populations.

Erector spinae plane block (ESPB) has emerged as a particularly attractive alternative within multimodal, opioid-sparing programs. ESPB is an ultrasound-guided interfascial technique in which local anesthetic is deposited deep to the erector spinae muscle over the thoracic transverse processes, with cranio-caudal and anterior spread that can reach the paravertebral region and produce multilevel dorsal and ventral rami blockade [6]. Several features recommend ESPB for the sternotomy context: the sono-anatomy is consistent and the target is superficial and compressible; the injection site is anatomically distant from the neuraxis and major vessels; and the block can be performed safely in patients receiving anticoagulation typical of cardiac surgery pathways [7]. Mechanistically, the capacity for wide dermatomal coverage aligns with the diffuse nociceptive inputs generated by midline sternotomy and mediastinal drains. Clinically, early randomized and comparative investigations have suggested reductions in perioperative opioid consumption, improvements in pain intensity scores, facilitation of earlier ventilatory weaning, and lower rates of postoperative nausea and vomiting when ESPB complements standardized systemic regimens [8]. Continuous catheter techniques further allow programmed redosing beyond the expected duration of single-shot injections, potentially sustaining analgesia through the period of peak post-sternotomy pain. At the same time, ESPB’s complication profile appears favorable relative to neuraxial techniques, with rare events (e.g., pneumothorax, local anesthetic systemic toxicity) that can be mitigated through ultrasound guidance, weight-based dosing, and structured monitoring [9].

Despite these promising signals, the aggregate literature remains heterogeneous and key questions are unresolved. Two recent reviews frame the current uncertainty. First, a broad synthesis that emphasized pain-related endpoints reported improvements in early postoperative pain and opioid-related outcomes with single-shot ESPB but did not resolve the question of comparative safety, largely because block-related adverse events were not prespecified as primary outcomes [10]. Second, the most recent 2025 meta-analysis restricted to adult randomized controlled trials found no difference in 24-hour coughing pain (its primary endpoint) yet demonstrated lower coughing and resting pain at 48–72 hours, reduced 24-hour opioid consumption, and shorter mechanical-ventilation duration with ESPB; it detected no differences in ICU or hospital length of stay and graded the overall certainty as moderate-to-low due to substantial clinical and statistical heterogeneity [11]. Importantly, between-study variation in surgical indications, anesthetic co-interventions, block timing and volumes, and—crucially—technique (single-shot versus catheter-based continuous infusion) complicates pooled effect estimates and limits bedside generalizability. Consequently, while efficacy signals exist, decision-makers still lack a technique-specific, adult-only, sternotomy-focused synthesis that treats safety as co-primary with efficacy.

Our systematic review and meta-analysis, is designed to solve this problem. We will focus exclusively on adults undergoing conventional cardiac procedures (e.g., coronary artery bypass grafting and valve surgery via median sternotomy) to reduce clinical heterogeneity. We will predefine technique-based subgroups that contrast single-shot with catheter-based ESPB, enabling a direct test of whether continuous infusion confers incremental benefit over single injections. Outcomes will be standardized across clinically coherent time points: pain at 0, 24, 36, and 72 hours; cumulative opioid consumption; time to rescue analgesia; duration of mechanical ventilation; and ICU and hospital length of stay. Critically, safety will be elevated to a co-primary domain, with systematic capture of block-related adverse events including local anesthetic systemic toxicity, pneumothorax, vascular puncture, bleeding or hematoma, infection, and block failure. By integrating the latest efficacy evidence—including the 2025 randomized-trials meta-analysis—and explicitly addressing the safety blind spot of prior pain-focused syntheses, our work aims to deliver decision-grade estimates of both benefit and risk. We hypothesize that ESPB, when implemented within standardized perioperative pathways, will reduce early opioid exposure and improve multidimensional recovery while maintaining a favorable safety profile relative to neuraxial and systemic-only strategies—providing the clarity necessary to guide guideline development and routine adoption in adult cardiac surgery.

## 2. Methods

### 2.1 Literature Search

This meta-analysis was conducted according to a prespecified protocol and reported in accordance with PRISMA guidelines [12]. The Search was conducted using comprehensive databases like PubMed, Scopus, Embase, and CENTRAL till August 2025. The study protocol is registered with PROSPERO (International Prospective Register of Systematic Reviews) under registration number CRD420251124891. The search targeted studies meeting prespecified eligibility criteria—evaluations of the analgesic efficacy of erector spinae plane block (ESPB) in cardiac surgery. Guided by the PICOS framework, we used the Boolean expression “erector spinae plane block” AND “cardiac surgery.” Reference lists of eligible and pertinent articles were hand-searched to identify additional records. No language limits were applied. Full search string is in Supplementary File.

### 2.2 Screening

All citations retrieved from the four databases were exported into Rayyan, a web-based platform for screening and de-duplication, where duplicates were identified and removed. We then applied a priori eligibility criteria for quantitative synthesis: (i) peer-reviewed, full-text publications in English; (ii) exclusion of case reports, protocols, letters, narrative/systematic reviews and meta-analyses, conference abstracts, ongoing/unpublished studies, and observational designs; (iii) randomized controlled trials with complete outcomes; (iv) intervention arms evaluating ESPB in the context of cardiac surgery; (v) comparator arms using another regional technique, sham/placebo, or no block; and (vi) reporting of pain outcomes suitable for pooling. The primary outcome was coughing pain at 24 hours after surgery. Secondary outcomes comprised: (1) coughing and resting pain at additional postoperative time points; (2) 24-hour opioid consumption (standardized to morphine equivalents when necessary); (3) time to first rescue analgesic; (4) perioperative time variables—durations of surgery, anesthesia, mechanical ventilation, cardiopulmonary bypass, and aortic cross-clamp; and (5) lengths of stay in the intensive care unit and the hospital.

### 2.3 Data Extraction and Statistical Analysis

Two reviewers independently extracted data into Microsoft Excel 2021; discrepancies were resolved by consensus. Variables were grouped into three domains: (i) demographics (publication year, first author, sample size, age range); (ii) surgical characteristics (intervention/comparator details, operative classification); and (iii) efficacy metrics (pain by VAS/NRS, postoperative opioid use converted to morphine milligram equivalents, local-anesthetic dosing). The primary endpoint was coughing pain at 24 hours. Continuous outcomes were pooled as mean differences (MD) with 95% confidence intervals using Mantel–Haenszel random-effects models. When medians and interquartile ranges were reported, values were converted to means and standard deviations using standard methods. Heterogeneity was quantified with I²; random-effects models were used for I²>50%, otherwise fixed-effects were applied. Prespecified subgroup analyses compared continuous versus single-shot ESPB. Publication bias was assessed with funnel-plot symmetry. Statistical significance was set at p<0.05. Analyses were performed in Stata 18.0.

### 2.4 Risk of Bias Assessment

Two investigators independently assessed risk of bias using the Cochrane Risk of Bias 2 (RoB 2) tool for randomized trials. Certainty of evidence for each prespecified outcome was appraised with the GRADE framework, considering risk of bias, inconsistency, indirectness, imprecision, and publication bias, and categorized as high, moderate, or low.

## 3. Results

### 3.1 Baseline Characteristics

The selection of studies is summarized systematically using a PRISMA flowchart (Figure 1). The total number of records screened were 2658, out of which 24 Randomized Control Trial were included [13–36]. Geographically, India has a greater number of studies, number to be 8 [13,22,24,25,26,28,32,33] China 3 [14,17,18], Egypt 3 [27,30,31], Turkey 3[15,29,36], Belgium 2 [34,35], Romania 2 [20,21], Poland 1 [16], South Korea 1 [23] and Vietnam 1 [19]. Total number of patients were 1687, out of which 1073 were male, 637 were female, average age was 55.72 ± 6.40 with average follow up hours were 44.75 ± 18.29. Total number of patients in treatment group were 856 and total number of patients in control group were 837. The characteristics of each study is in Table S1.

**Figure 1.**
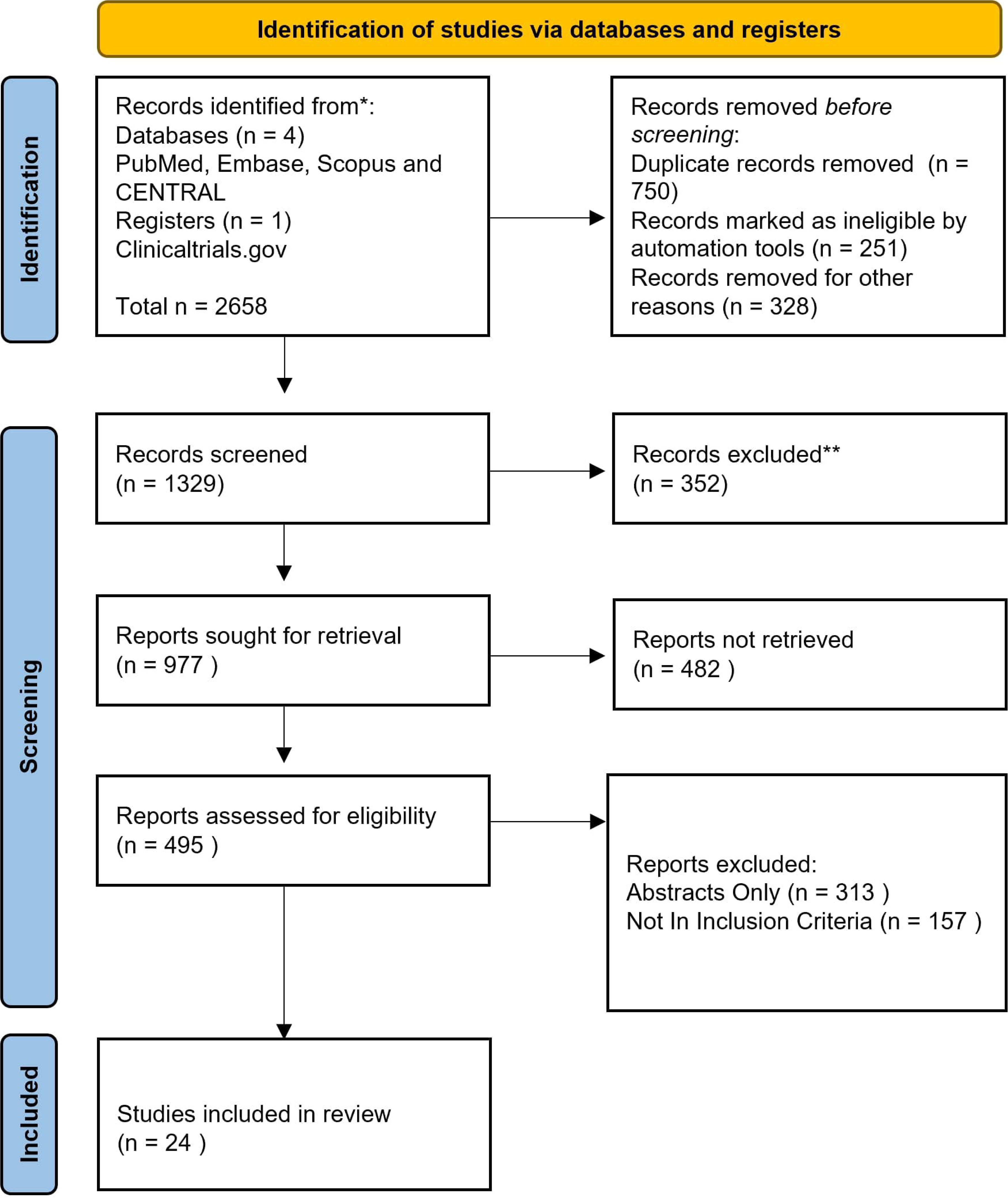
PRISMA Flow Diagram.

### 3.2 Postoperative Pain Scores at Rest

Across studies reporting resting VAS pain scores post-operation, the pooled effects were as follows: at 0 h, 20 studies [13–19,21,23,25–27,30–35], yielded a mean difference (MD) of −0.03 (95% CI −0.36 to 0.30) (Figure 2), a null effect that nominally favored the intervention; at 24 h, 20 studies [13–19,21,23,25–27,30–35], showed MD 0.01 (95% CI −0.32 to 0.34) (Figure 3), again indicating no difference; at 36 h, 9 studies [13,14,23,25,26,27,29,30,34] showed MD 0.50 (95% CI 0.08 to 0.92) (Figure 4), reflecting a small but statistically significant difference (interpreted as higher pain in the treatment arm if MD = treatment − control); and at 72 h, 3 studies [13,18,19] showed MD 0.27 (95% CI −0.40 to 0.94) (Figure 5), indicating no statistically significant difference.

**Figure 2.**
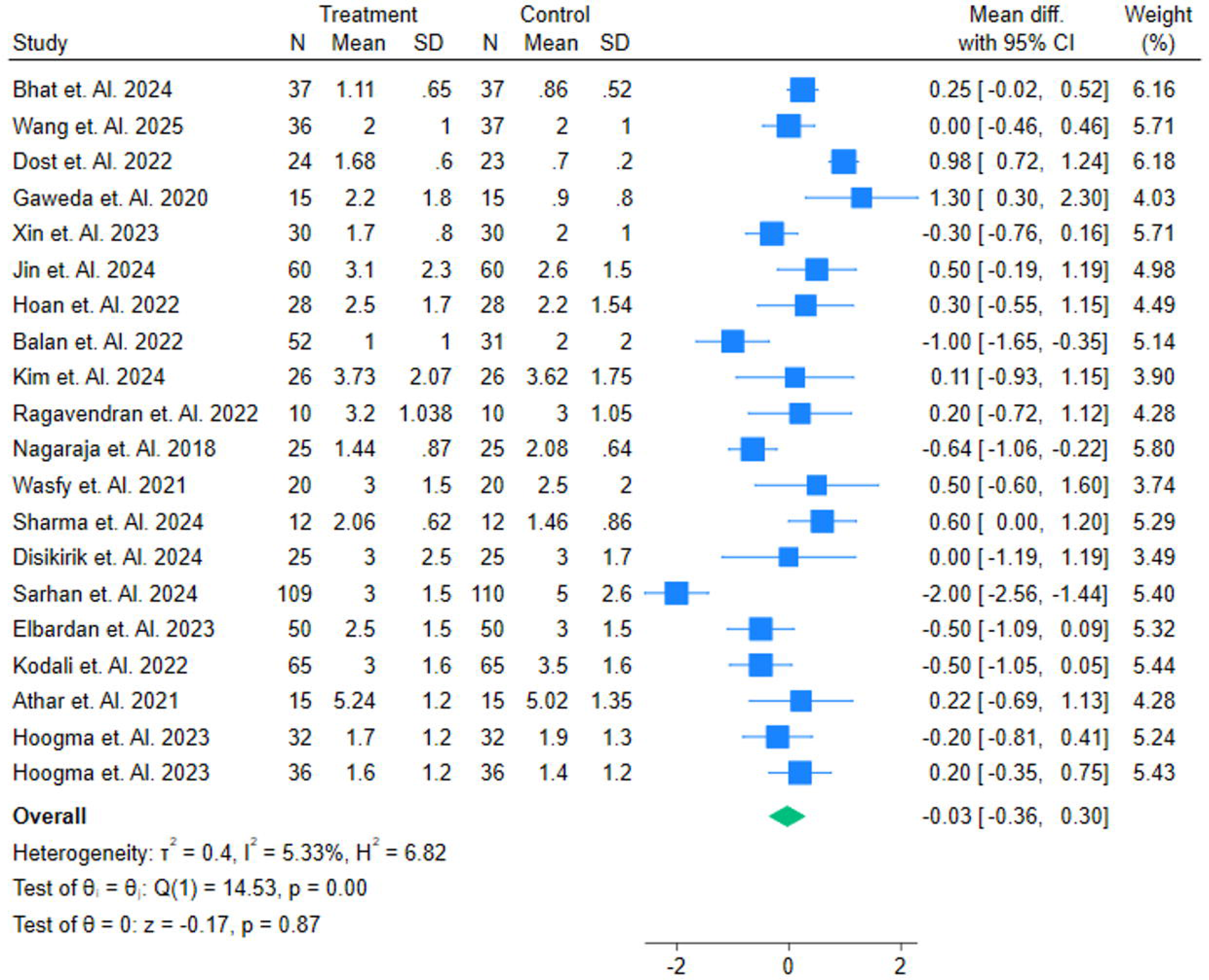
Pain Scores while at rest at O hours Post-Operation, evaluating the efficacy with Mean Difference with Mean ± SD.

**Figure 3.**
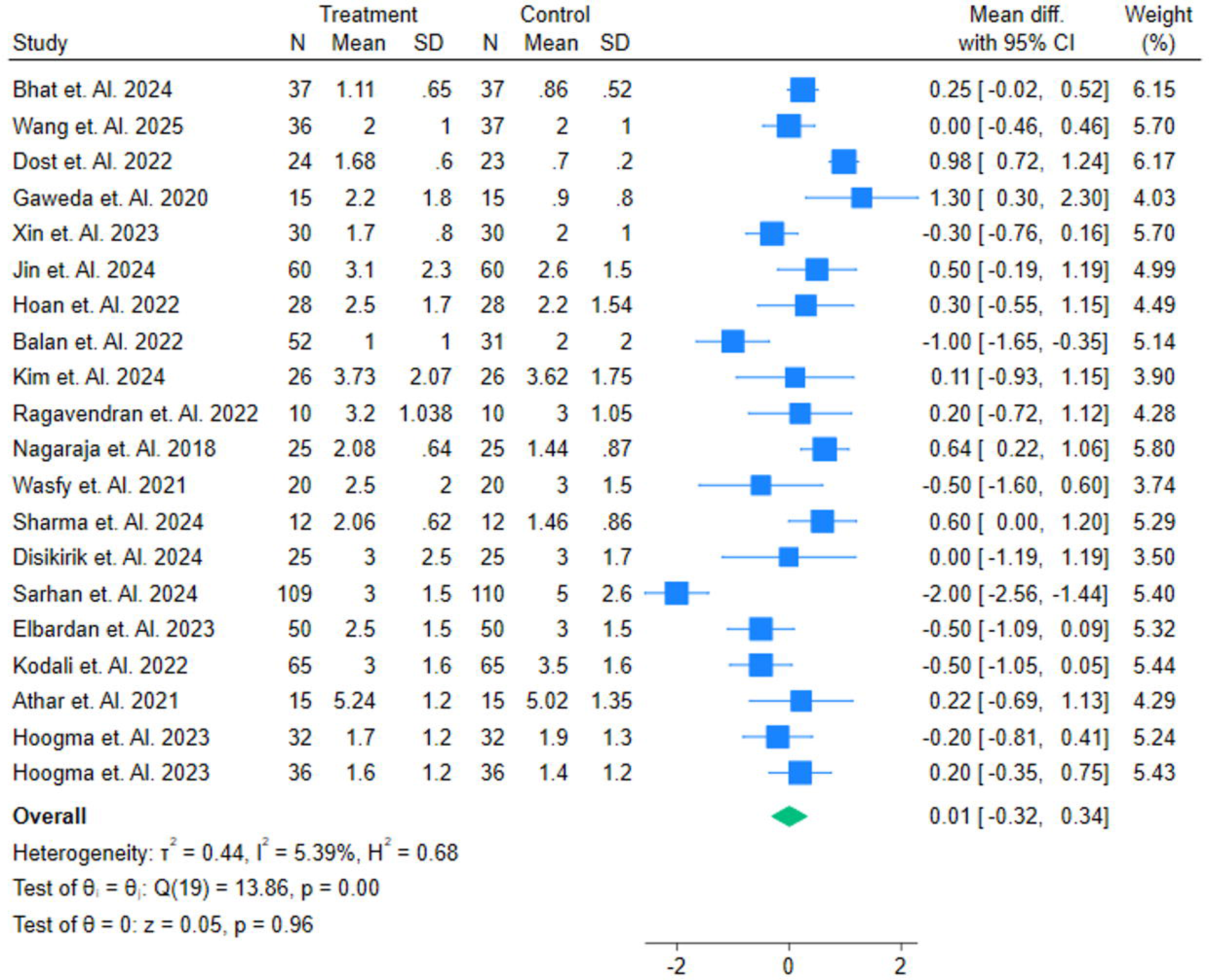
Pain Scores while at rest at 24 hours Post-Operation, evaluating the efficacy with Mean Difference with Mean ± SD.

**Figure 4.**
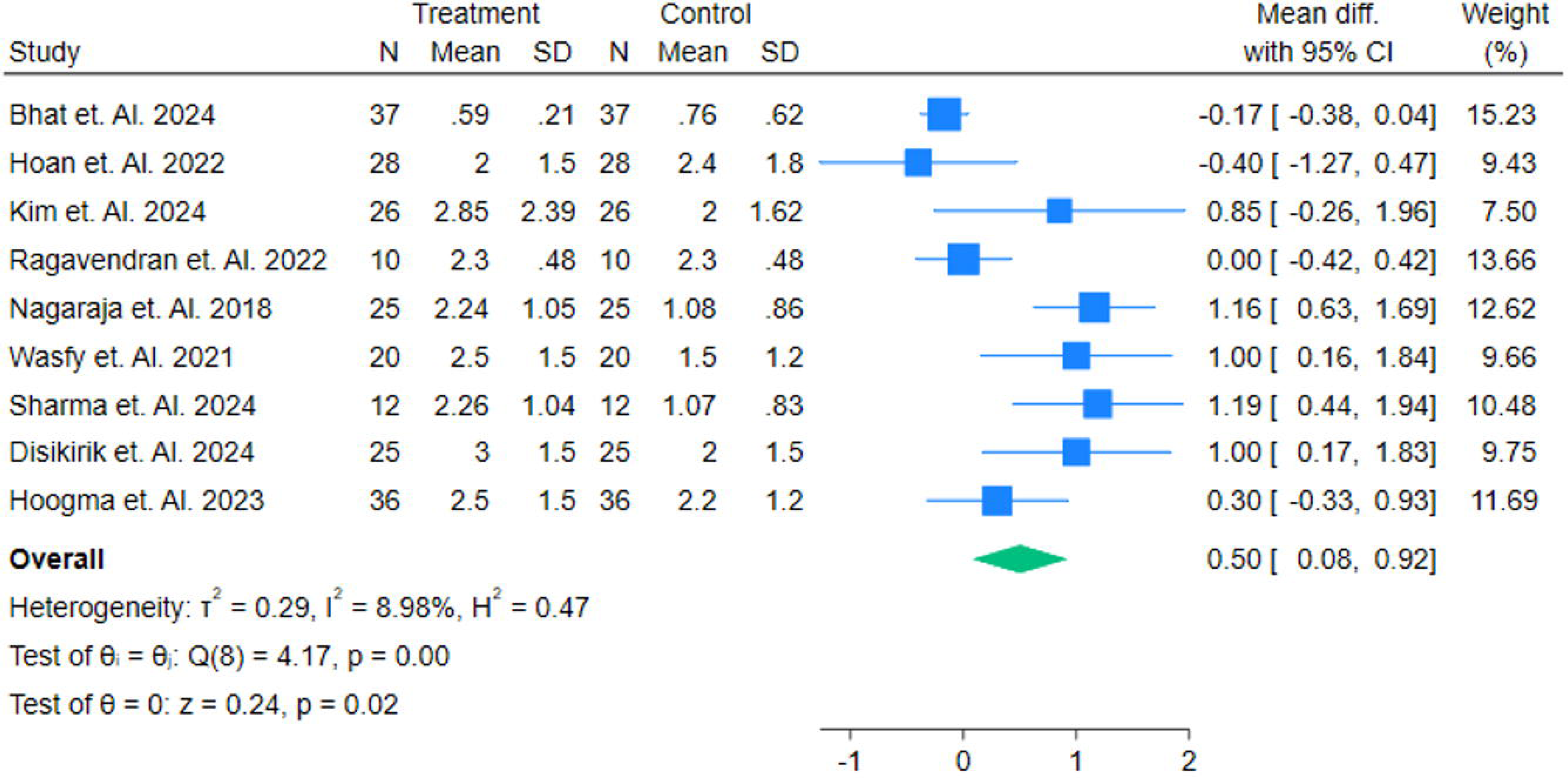
Pain Scores while at rest at 36 hours Post-Operation, evaluating the efficacy with Mean Difference with Mean ± SD.

**Figure 5.**
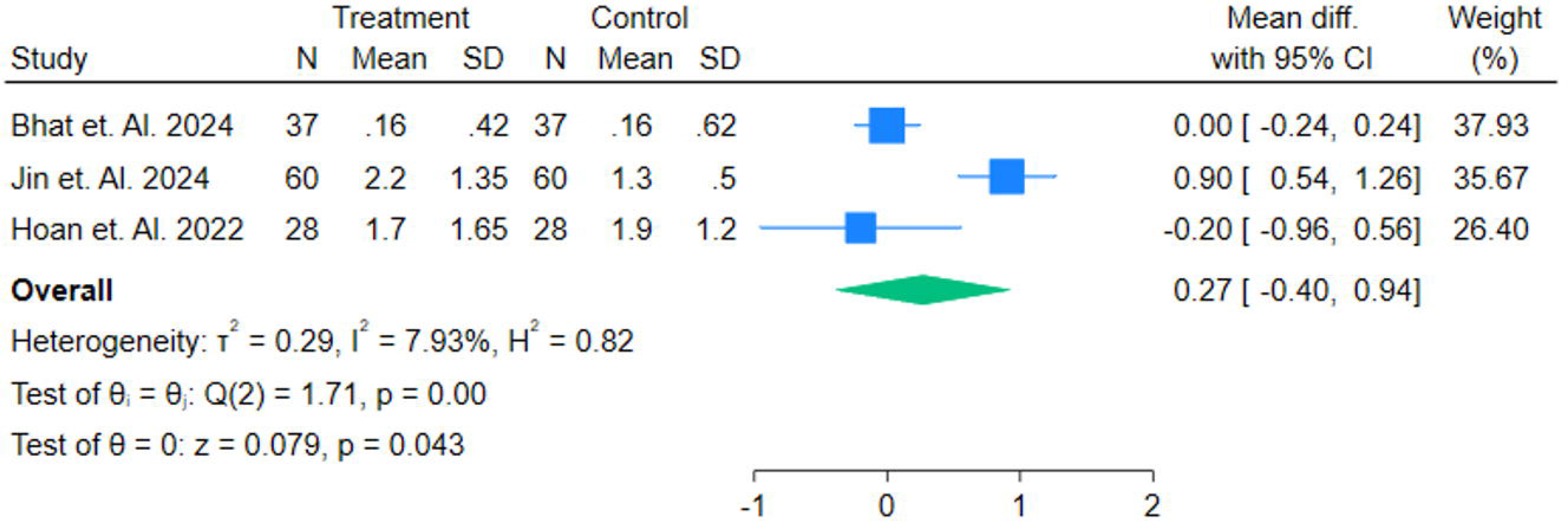
Pain Scores while at rest at 72 hours Post-Operation, evaluating the efficacy with Mean Difference with Mean ± SD.

### 3.3 Postoperative Pain Scores while coughing

Pooled analyses of post-operative VAS pain scores yielded the following mean differences (MD) with 95% confidence intervals: at 0 h, MD = 0.76 (0.03–1.48; 6 studies) (Figure 6) [13,15,18,25,26,28]; at 24 h, MD = 1.12 (0.03–2.21; 5 studies [13,18,25,26,28]); at 36 h, MD = 0.74 (0.06–1.43; 4 studies [13,25,26,28]) (Figure 7); and at 72 h, MD = 0.25 (−0.82–1.32; 2 studies [13,18]) (Figure 8). Estimates at 0, 24, and 36 h were statistically significant (CIs excluding zero), whereas the 72-h estimate was not.

**Figure 6.**
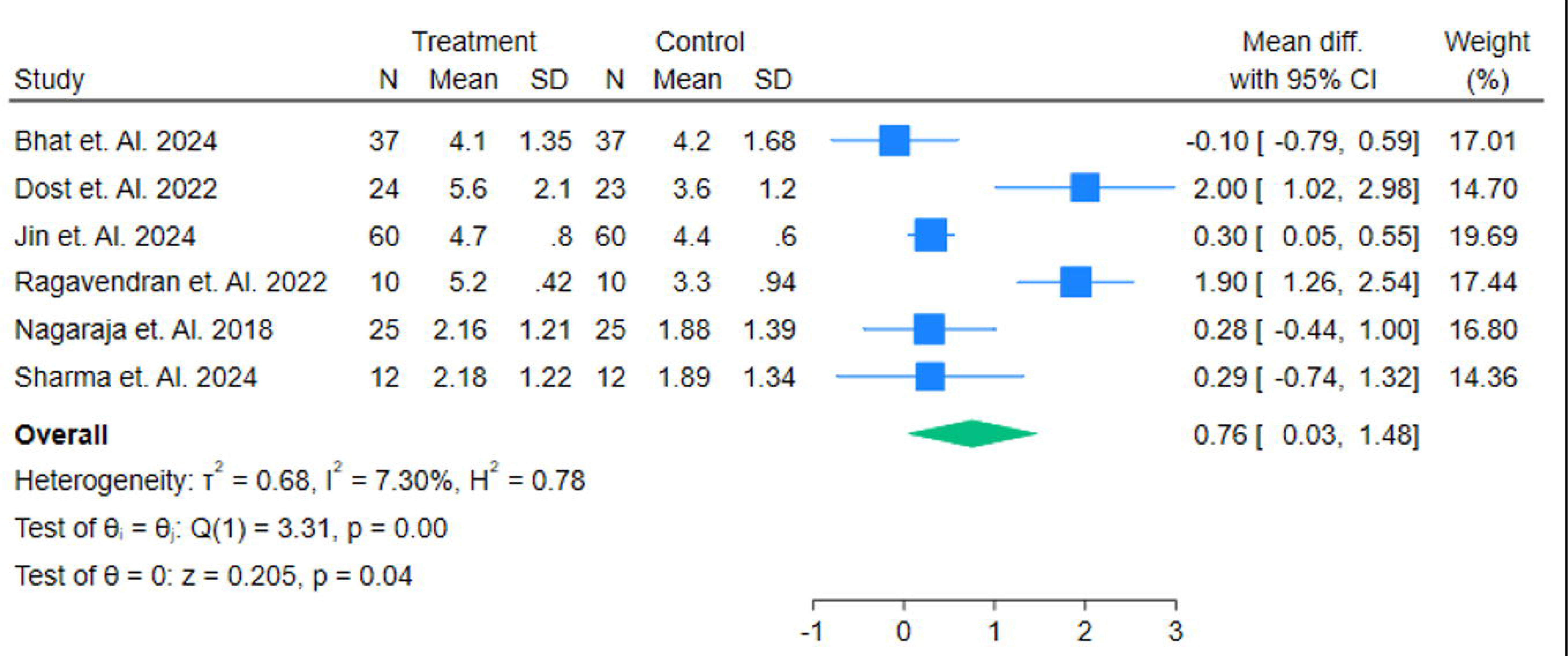
Pain Scores while Coughing at O hours Post-Operation, evaluating the efficacy with Mean Difference with Mean ± SD.

**Figure 7.**
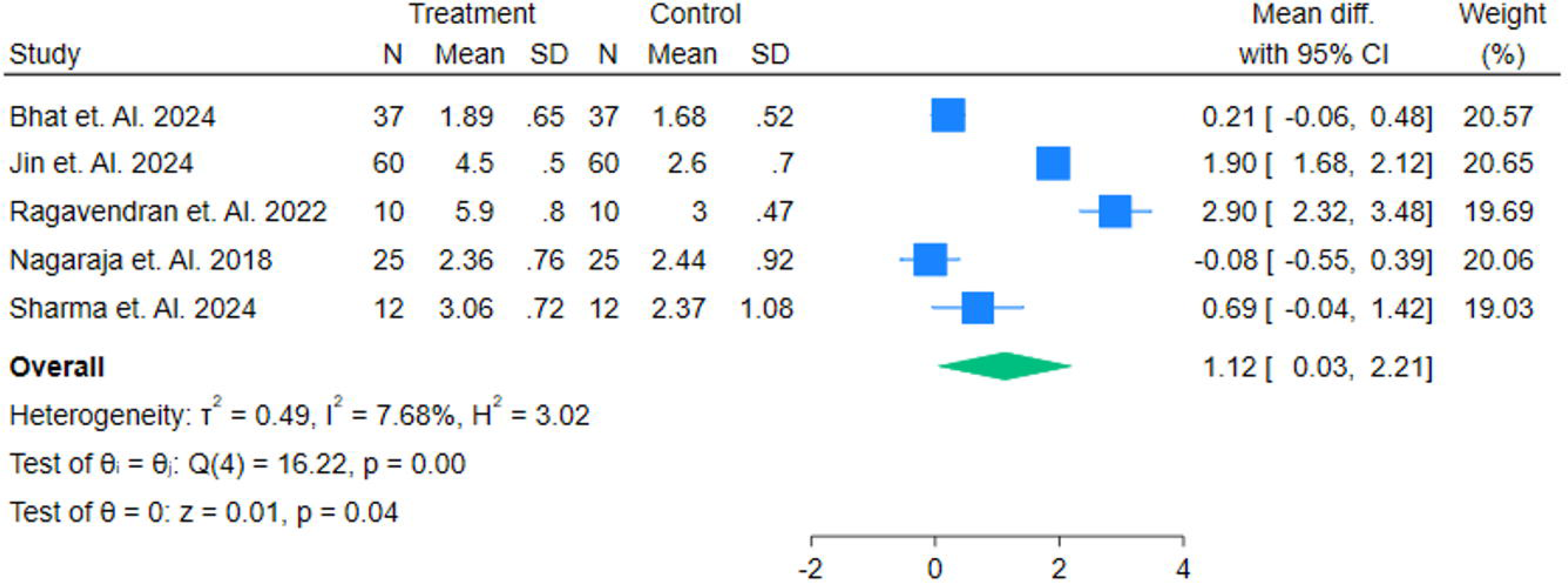
Pain Scores while Coughing at 24 hours Post-Operation, evaluating the efficacy with Mean Difference with Mean ± SD.

**Figure 8.**
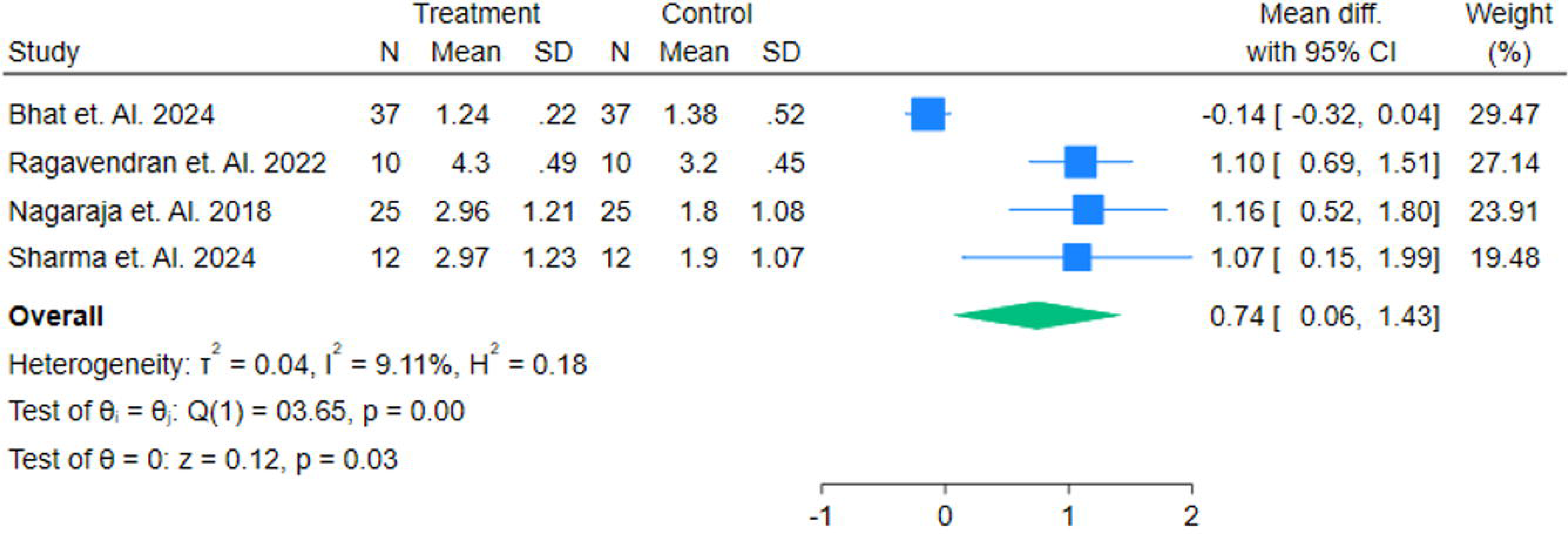
Pain Scores while coughing at 36 hours Post-Operation, evaluating the efficacy with Mean Difference with Mean ± SD.

### 3.4 Opioid Consumption post-operation

Opioid consumption (morphine-equivalent; mean difference, treatment − control; negative favors the intervention) showed a non-significant reduction at 0 h post-operation (MD −19.33; 95% CI −59.95 to 21.28) [16,20,27,29,30,34] (Figure 9) and at 24 h (MD −6.62; 95% CI −16.46 to 3.23) [16,17,19,20,29](Figure 10), followed by a statistically significant reduction at 36 h (MD −24.87; 95% CI −34.27 to −15.48) [19,29] (Figure 11), indicating a clear opioid-sparing effect only at the 36-hour time point while earlier estimates were imprecise and compatible with no difference.

**Figure 9.**
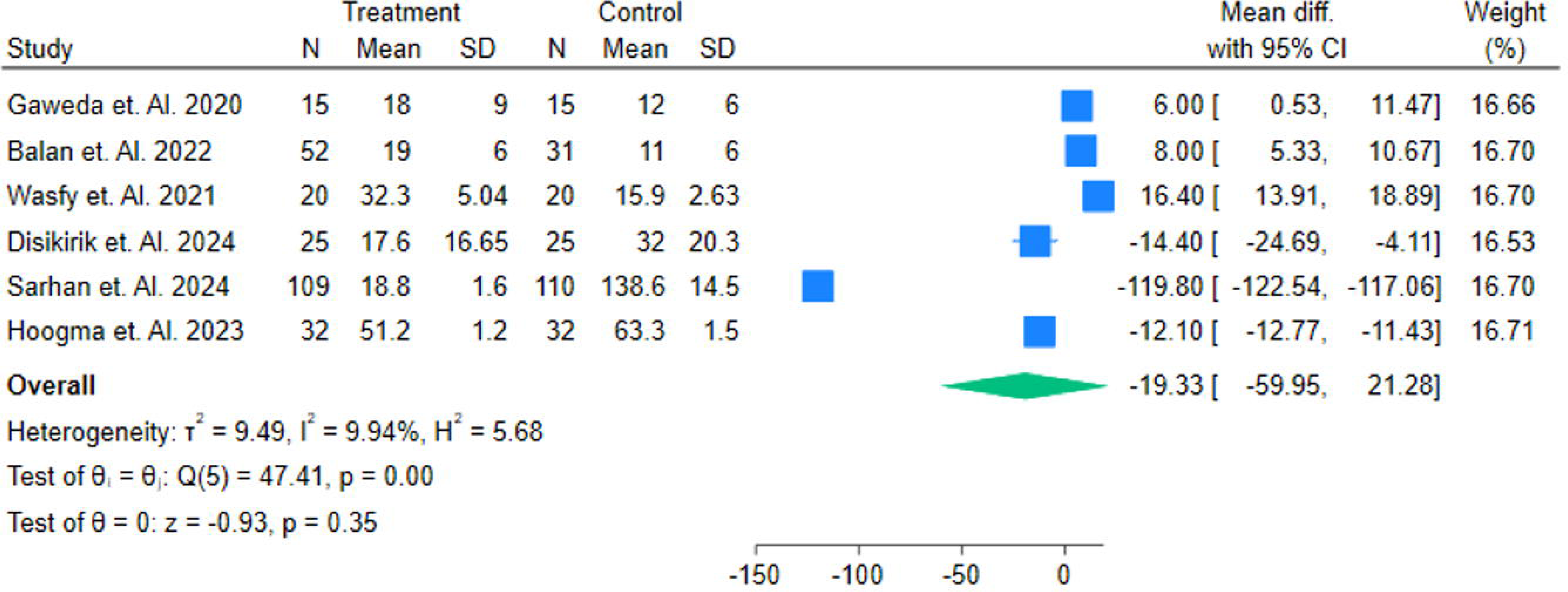
Forest plot of postoperative opioid consumption at Oh (morphineequivalent); pooled mean difference in Mean± SD.

**Figure 10.**
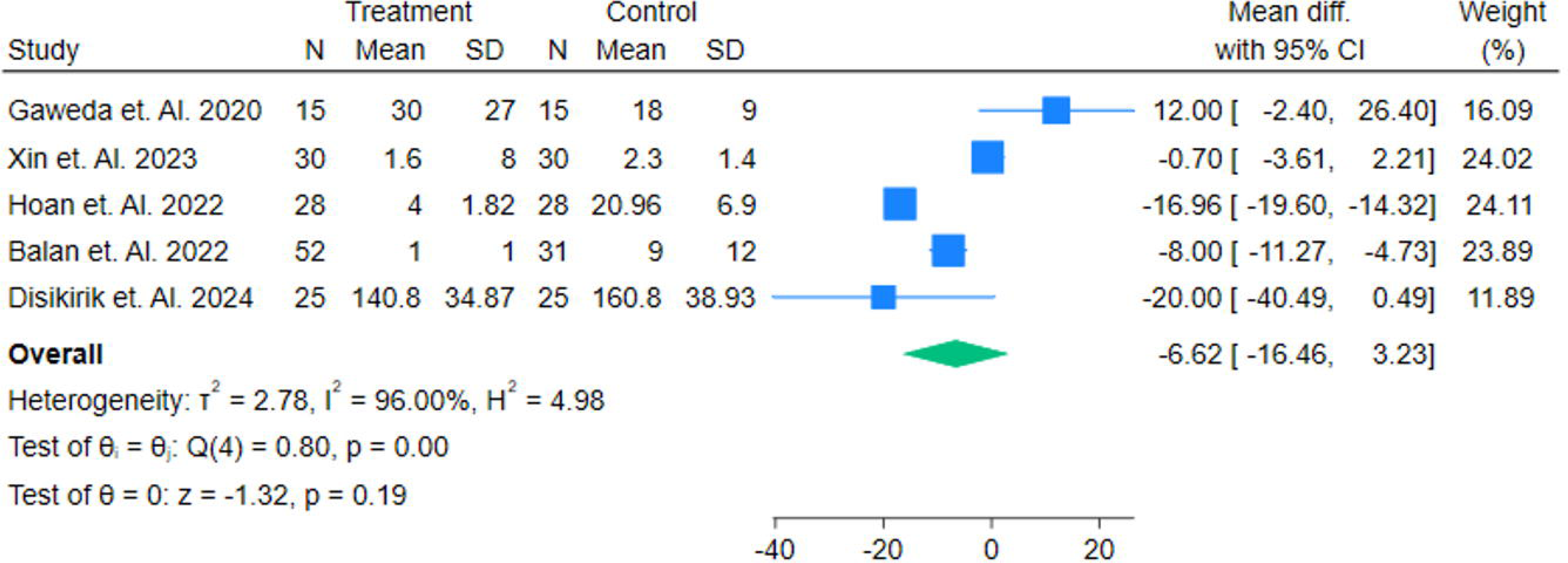
Forest plot of postoperative opioid consumption at 24 h (morphineequivalent); pooled mean difference in Mean± SD.

**Figure 11.**
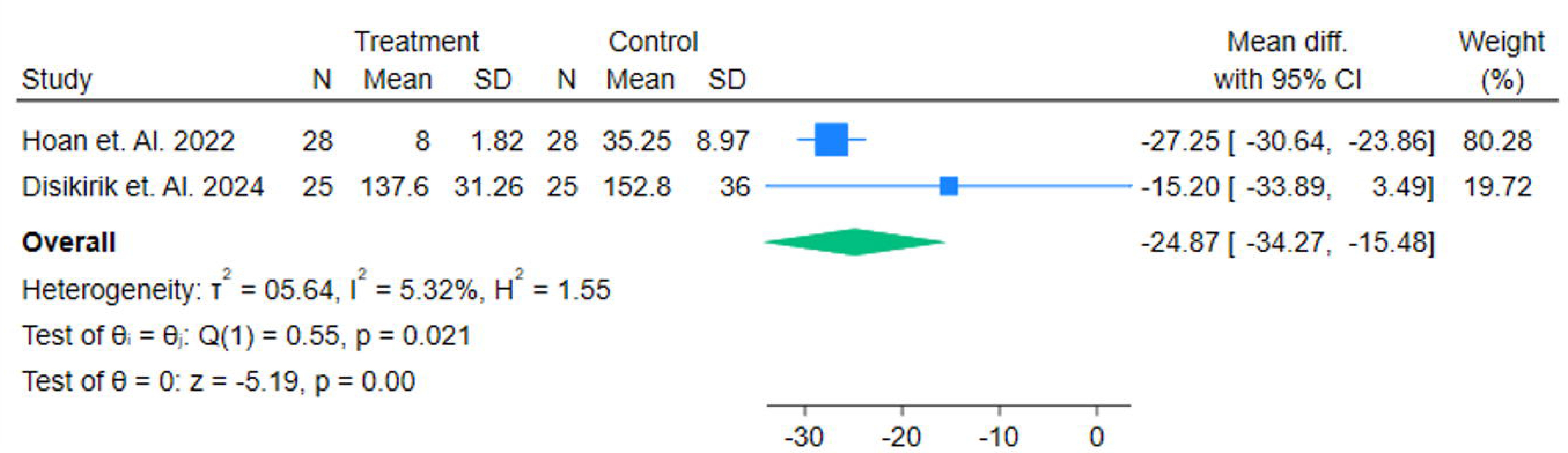
Forest plot of postoperative opioid consumption at 36 h (morphineequivalent); pooled mean difference in Mean± SD.

### 3.5 Length of Stay

Sixteen studies [13,14,17,19,20,23,24,25,26,27,31,32,33–35] reported length of stay. The pooled mean difference (MD) was −0.21 (95% CI, −0.70 to 0.28) (Figure 12) days, indicating Treatment group has significant impact in reducing LOS between intervention and control groups.

**Figure 12.**
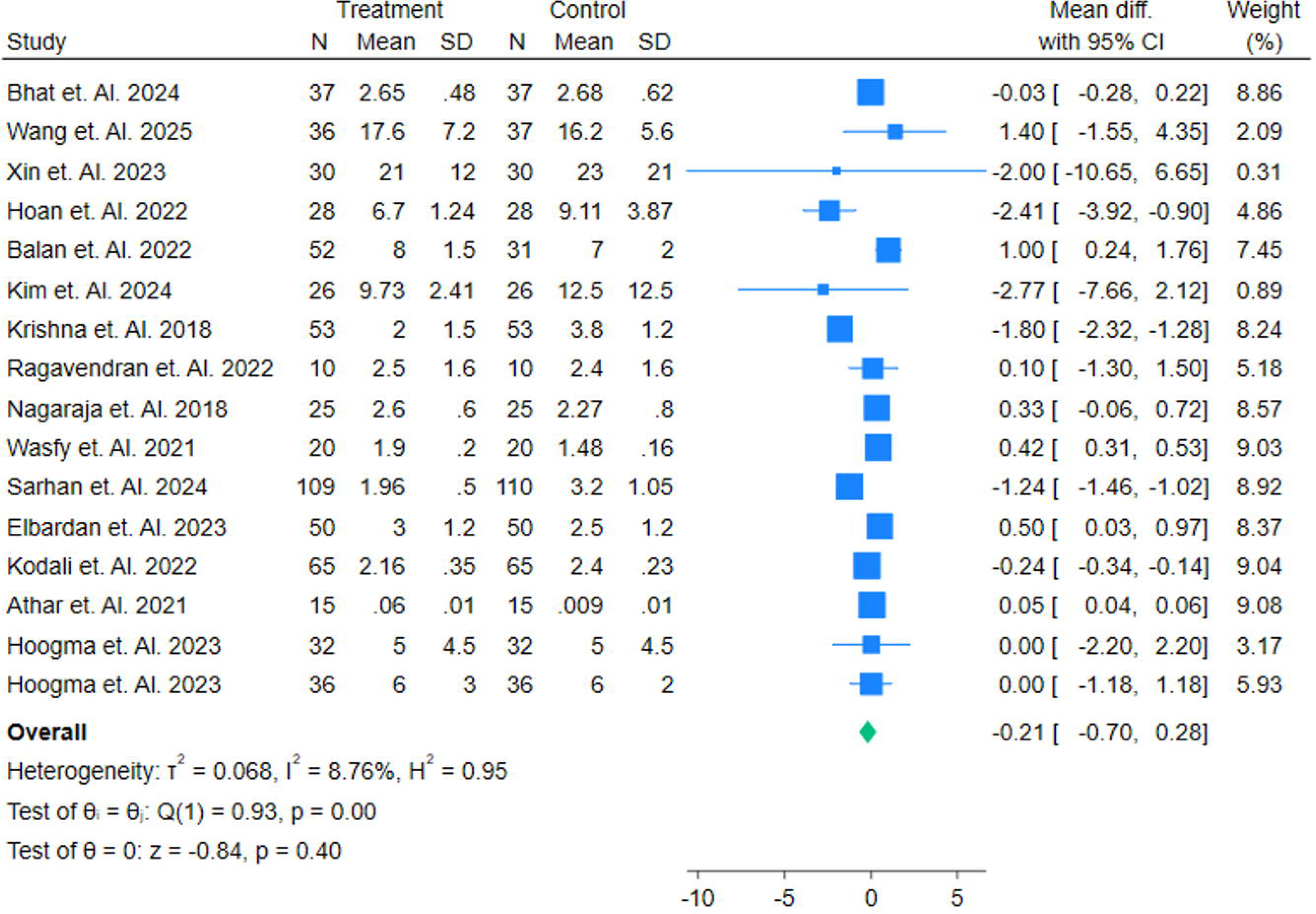
Forest plot of hospital length of stay (days); pooled mean difference in Mean± SD.

### 3.6 Adverse Events

Across safety endpoints, point estimates generally favoured the treatment arm, although precision was limited and all 95% confidence intervals spanned the null. Arrhythmia was reported in seven studies [13,14,18,20,23,34,35] (log risk ratio [log RR] 0.12; 95% CI −0.32 to 0.55) (Figure 13). Injection-site reactions were reported in three studies [18,30,34] (log RR 0.35; 95% CI −2.17 to 2.87) (Figure 14). Neurological complications were reported in three studies (log RR 0.47; 95% CI −0.43 to 1.37) (Figure 15). Postoperative nausea and vomiting (PONV) were reported in eight studies [14,17–20,33–35] (log RR −0.39; 95% CI −0.86 to 0.08) (Figure 16). Taken together, these findings indicate no statistically significant differences between groups for these adverse outcomes, with point estimates tending to favour the treatment for safety profile. Grade quality of all articles was High (Table S1) and ROBS 2.0 was low (Figure S1).

**Figure 13.**
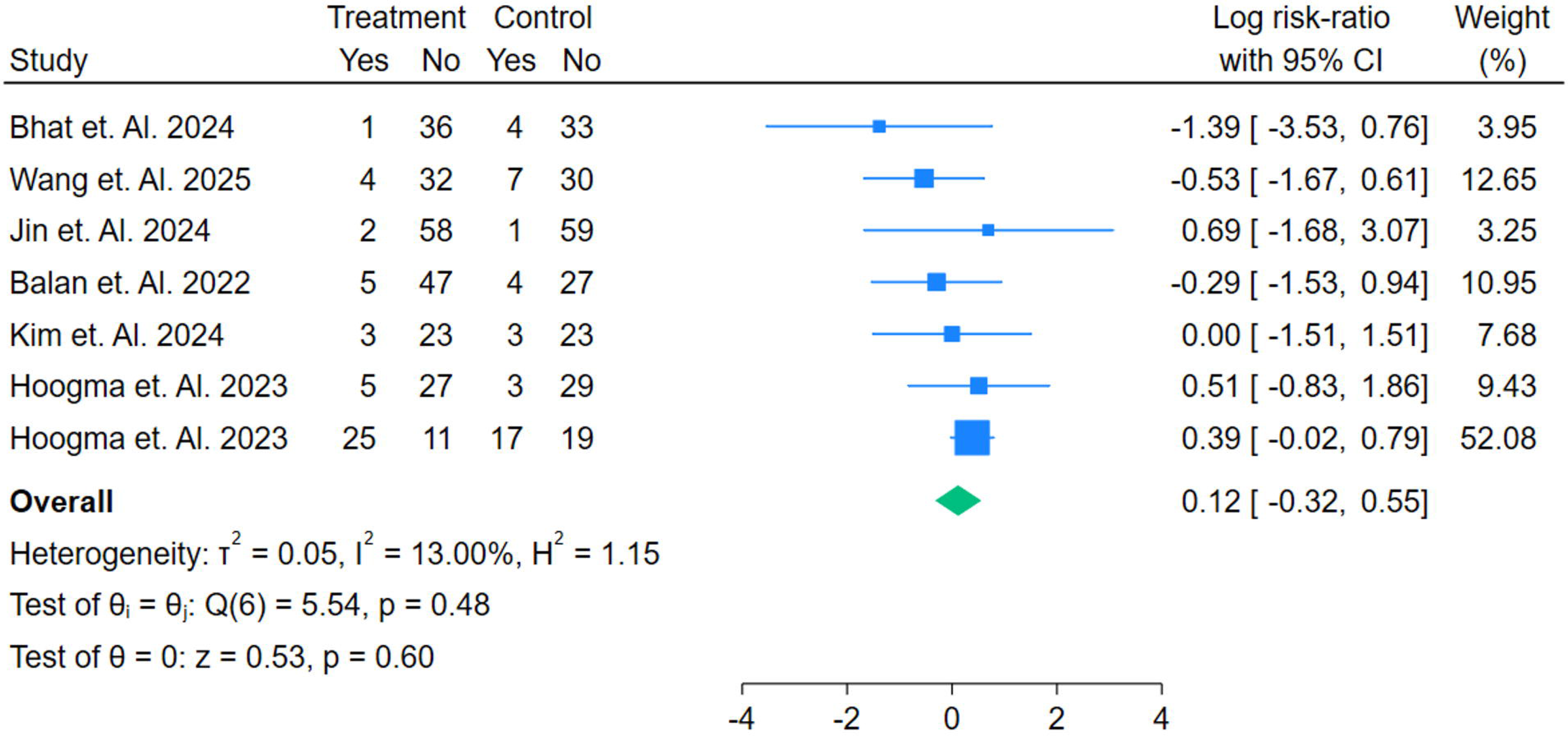
Forest plot of Adverse events of Arrhythmia in Risk Ratios.

**Figure 14.**
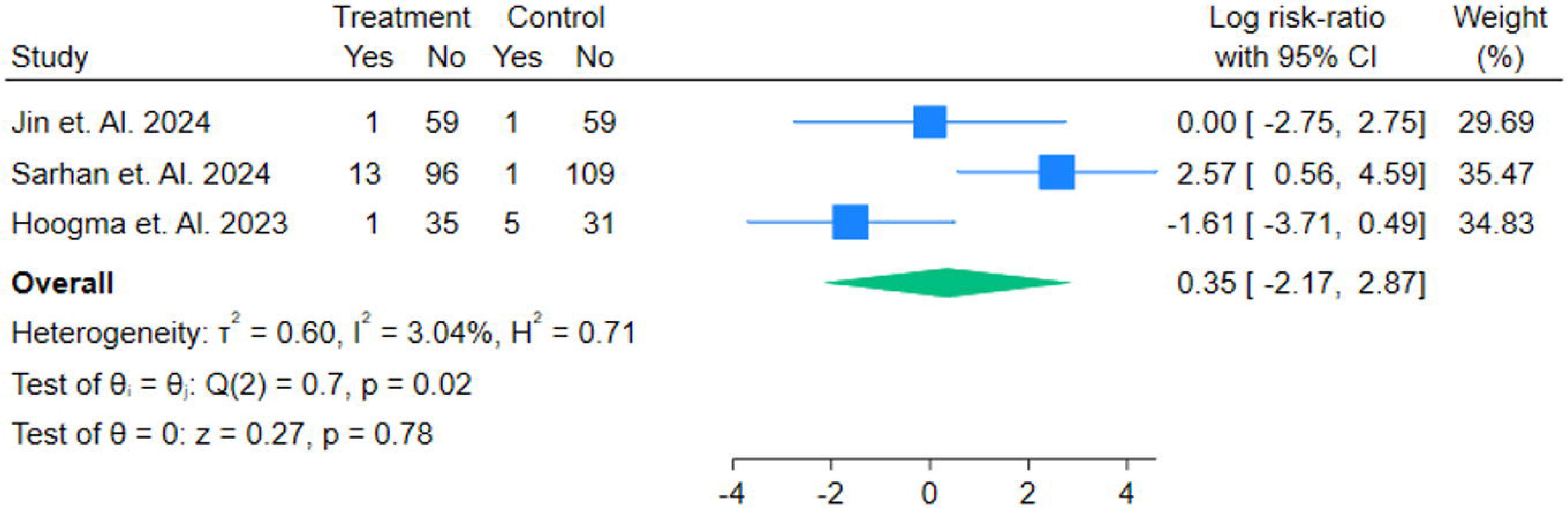
Forest plot of Adverse events of Injection Site Reaction in Risk Ratios.

**Figure 15.**
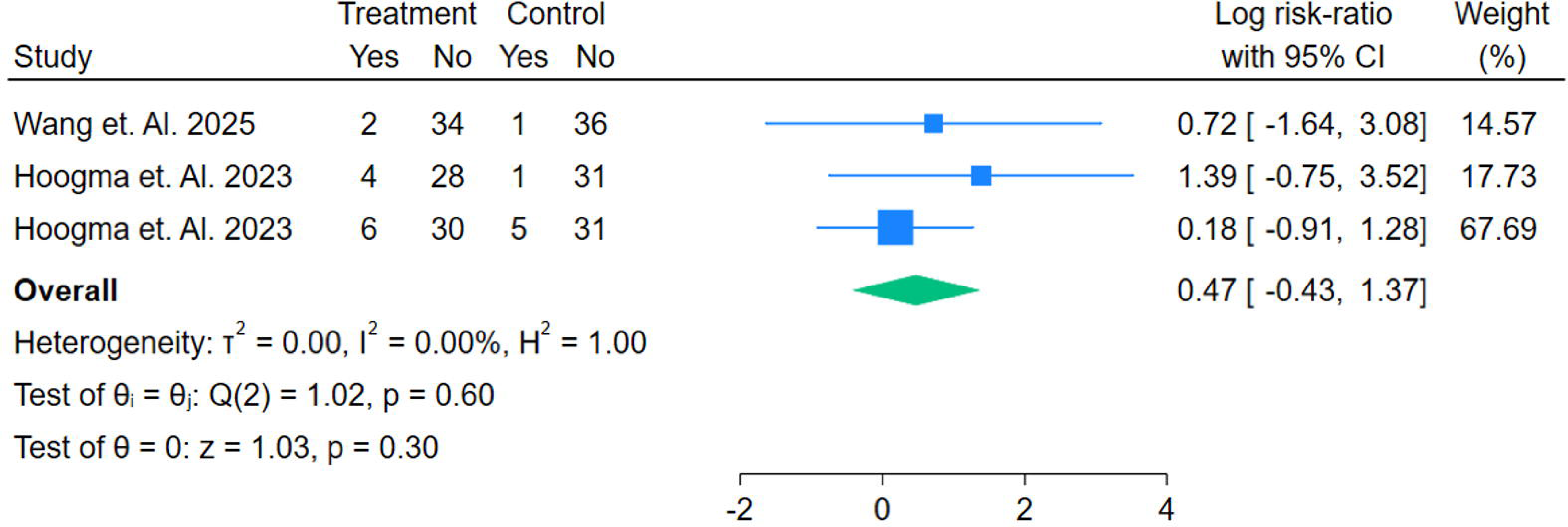
Forest plot of Adverse events of Neurological Consequences in Risk Ratios.

**Figure 16.**
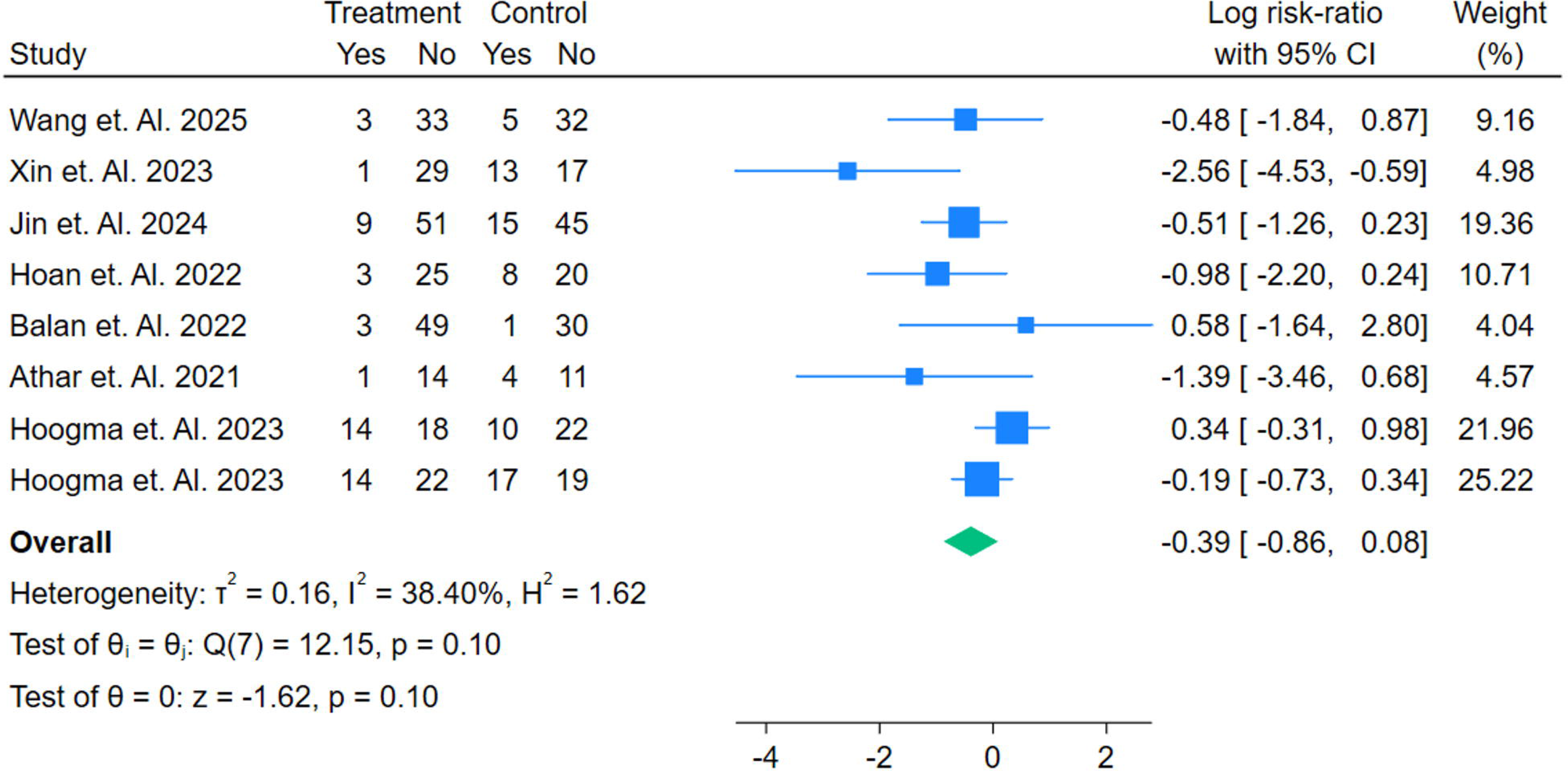
Forest plot of Adverse events of Post Operative Nausea and Vomiting in Risk Ratios.

## 4. Discussion

The present synthesis aligns with, yet also nuances, the evolving evidence bases on erector spinae plane block (ESPB) after adult median sternotomy. Several meta-analyses now converge on an opioid-sparing signal and improvements in selected recovery metrics, albeit with nontrivial heterogeneity. For example, a 2024 systematic review restricted largely to single-shot ESPB reported reductions in early pain (4 h), cumulative postoperative opioid use, mechanical-ventilation duration, and postoperative nausea and vomiting, with no clear effect on ICU or hospital length of stay—findings broadly consonant with our pooled estimates for ventilatory and opioid outcomes but not for length-of-stay endpoints [37]. By contrast, an earlier review that included mixed designs and broader cardiothoracic populations suggested benefits spanning intraoperative and 48-h opioid consumption, pain scores, extubation timing, and both ICU and hospital stay; differences in eligibility (e.g., pediatric inclusion, non-sternotomy procedures) and cointerventions likely explain the more expansive effect profile there [38]. The most recent adult RCT–focused meta-analysis (2025) further refines these signals. It found no difference in 24-h coughing pain (its primary endpoint) but did demonstrate lower coughing and resting pain at 48–72 h, reduced 24-h morphine consumption, and shorter mechanical-ventilation duration; length-of-stay effects were neutral, and certainty was downgraded for inconsistency and imprecision [11]. This pattern— attenuated early analgesic advantage with benefits manifesting later and in resource-use measures—mirrors the time-course we observed for pain at rest and opioid outcomes. Similarly, an adult-only RCT meta-analysis from 2024 concluded ESPB reduced pain within 12 h and decreased 24-h opioid use, while emphasizing very-low-to-moderate certainty and the need for standardization—again consistent with our heterogeneity estimates and the variability we recorded in local-anesthetic dosing, volumes, timing, and bilateral versus unilateral approaches [39]. Technique matters. Single-shot versus catheter-based continuous ESPB remains a key uncertainty. Preliminary meta-analytic work in minimally invasive cardiac surgery suggests continuous infusion may shorten hospital stay, hinting at a dose–duration relationship that single injections cannot reliably achieve; however, estimates are imprecise and drawn from limited RCTs. Our prespecified subgrouping by technique directly addresses this gap and should help clarify whether continuous regimens deliver incremental benefit. Outside the cardiac domain, comparisons of ESPB with thoracic paravertebral block in thoracic surgery often show broadly similar analgesia with favorable safety for ESPB, albeit with debates over injectate spread and dermatomal reliability—context that supports, but does not substitute for, sternotomy-specific trials.

Safety warrants equal attention. Neuraxial techniques such as thoracic epidural analgesia have long been considered effective after sternotomy, but anticoagulation and the rare yet severe risk of epidural hematoma continue to temper enthusiasm in routine cardiac pathways. These constraints motivate interest in fascial plane blocks that are anatomically distant from the neuraxis and more compressible—properties that may confer procedural safety advantages, even if high-quality head-to-head safety data remain sparse. Importantly, recent ESPB meta-analyses in cardiac surgery reported few block-related complications and no consistent safety signal against ESPB, though most trials were not powered primarily for adverse events—hence our decision to elevate block-specific harms (e.g., pneumothorax, local anesthetic systemic toxicity, bleeding/hematoma, infection, block failure) to a co-primary domain. Taken together, contemporary evidence supports ESPB as a practical component of multimodal, opioid-sparing analgesia after adult sternotomy, with the clearest and most consistent benefits in opioid exposure, late-phase pain, and ventilatory weaning rather than in length-of-stay endpoints. Differences across meta-analyses likely reflect study-level heterogeneity in surgical indications, ERAS cointerventions, block timing, local-anesthetic dose/volume, and, crucially, single-shot versus continuous techniques. By restricting to adult sternotomy, prespecifying technique-based subgroups, harmonizing time points, and treating safety as co-primary, the present review is positioned to deliver more decision-useful estimates while highlighting where targeted, adequately powered RCTs are still needed—particularly for continuous catheter strategies within standardized perioperative pathways.

## 5. Conclusion

ESPB is a practical, ultrasound-guided adjunct for adult sternotomy that consistently lowers opioid exposure— most clearly by 36 hours—while showing a low, comparable rate of adverse events. Effects on early resting or coughing pain are small and inconsistent, and length of stay appears unchanged. Substantial heterogeneity (indications, co-interventions, timing, injectate volume, unilateral versus bilateral placement, and single-shot versus catheter techniques) likely dilutes pooled estimates. In anticoagulated cardiac pathways where neuraxial options are limited, ESPB fits multimodal, opioid-sparing care. Priority next steps are adequately powered, technique-specific trials—especially continuous-catheter protocols with standardized dosing, bilateral placement, and prespecified safety and recovery endpoints after sternotomy.

## Data Availability

In Supplementary File

## Conflict of Interest

The authors certify that there is no conflict of interest with any financial organization regarding the material discussed in the manuscript.

## Funding

The authors report no involvement in the research by the sponsor that could have influenced the outcome of this work.

## Authors’ contributions

All authors contributed equally to the manuscript and read and approved the final version of the manuscript.

## Notes

### Competing Interest Statement

The authors have declared no competing interest.

### Clinical Protocols

https://www.crd.york.ac.uk/PROSPERO/view/CRD420251124891

### Funding Statement

None

