## Supplement for "Analgesic Efficacy and Perioperative Safety of the Erector Spinae Plane Block in Adult Cardiac Surgery"

Table S1. Demographic Characteristics of the Studies

| References | Author Name Year | Country | Total Sample | Male | Female | Mean Age | Mean Follow Up Hours | Treatment Group Name | Treatment Group Sample Size | Control Group Name | Control Group Sample Size | Main Finding | GRADE |
| --- | --- | --- | --- | --- | --- | --- | --- | --- | --- | --- | --- | --- | --- |
| 13 | Bhat et. Al. 2024 | India | 74 | 53 | 21 | 60.3 | 72 | Bilateral Erector Spinae Plane block | 37 | Thoracic Epidural Analgesia | 37 | The study concluded that the **erector spinae plane block (ESP)** offers an effective, safer alternative to thoracic epidural analgesia (TEA) in cardiac surgery, with better pain control, fewer complications, and improved recovery outcomes. | High |
| 14 | Wang et. Al. 2025 | China | 73 | 49 | 23 | 61.5 | 48 | Erector spinae plane block | 36 | Intercostal nerve block | 37 | No significant difference in postoperative analgesic effect between ESPB and ICNB in patients after MIDCAB. | High |
| 15 | Dost et. Al. 2022 | Turkey | 47 | 31 | 16 | 58.1 | 24 | Erector spinae plane block | 24 | Esp + S-PIP | 23 | The combination of ESP and S-PIP blocks led to reduced postoperative morphine consumption and improved pain scores in patients undergoing open cardiac surgery. | High |
| 16 | Gaweda et. Al. 2020 | Poland | 30 | 22 | 8 | 60.7 | 72 | Erector spinae plane block | 15 | PECS + ESP | 15 | The addition of PECS blocks to ESP block reduced opioid consumption, decreased pain intensity, and increased patient satisfaction with pain management after mitral/tricuspid valve repair via mini-thoracotomy. | High |
| 17 | Xin et. Al. 2023 | China | 60 | 47 | 13 | 60 | 48 | Erector spinae plane block | 30 | Saline | 30 | The study concluded that the **ESPB** provided **effective early postoperative analgesia**, reducing pain scores and opioid consumption compared to the control group. | High |
| 18 | Jin et. Al. 2024 | China | 120 | 65 | 55 | 54.1 | 144 | Erector spinae plane block | 60 | Saline | 60 | Continuous ESPB mildly improved the quality of recovery in patients undergoing minimally invasive cardiac surgery and reduced postoperative pain and opioid consumption without affecting chronic pain. | High |
| 19 | Hoan et. Al. 2022 | Vietnam | 46 | 23 | 23 | 41.5 | 72 | Erector spinae plane block | 28 | Patient Controlled Analgesia | 28 | ESPB provides better post-operative analgesia and reduces morphine requirements compared to PCA, with no significant complications | High |
| 20 | Balan et. Al. 2023 | Romania | 85 | 53 | 32 | 63 | 48 | Erector spinae plane block | 42 | Nociception | 43 | The addition of bilateral single-shot ESPB to general anesthesia improved the quality of intraoperative nociception control according to a NOL index-based evaluation. | High |
| 21 | Balan et. Al. 2022 | Romania | 83 | 43 | 40 | 63 | 48 | Erector spinae plane block | 52 | Saline | 31 | The study demonstrated that NOL index-directed bilateral ESPB significantly reduced intraoperative fentanyl consumption and postoperative morphine usage, facilitated faster extubation, and improved pain scores, without increasing adverse events compared to the control group. | High |
| 22 | Keshwani et. Al. 2023 | India | 38 | 24 | 14 | 36.6 | 24 | Pecto-Intercostal Fascial Block | 19 | Erector spinae plane block | 19 | PIFB and ESPB showed similar efficacy in postoperative analgesia and opioid consumption, with PIFB being more convenient in administration. | High |
| 23 | Kim et. Al. 2024 | South Korea | 56 | 52 | 10 | 64.5 | 48 | Erector spinae plane block | 26 | Control | 26 | ESPB reduces the incidence of severe pain and opioid use after OPCAB surgery. | High |
| 24 | Krishna et. Al. 2018 | India | 106 | 61 | 55 | 48.32 | 12 | Erector spinae plane block | 53 | Paracetamol and Tramadol | 53 | ESP block provided significantly better pain relief at rest for a longer duration compared to intravenous paracetamol and tramadol in adult cardiac surgery patients. | High |
| 25 | Ragavendran et. Al. 2022 | India | 20 | 20 | 0 | 52.5 | 48 | Erector spinae plane block | 10 | Thoracic Epidural Analgesia | 10 | Both ESP block and TEA provided comparable analgesia at rest, with no significant differences in post-surgery complications. | High |
| 26 | Nagaraja et. Al. 2018 | India | 50 | 25 | 25 | 50.12 | 54 | Thoracic Epidural Analgesia | 25 | Erector spinae plane block | 25 | ESP block is a promising alternative to TEA for optimal perioperative pain management in cardiac surgery, with comparable efficacy and a favorable safety profile. | High |
| 27 | Wasfy et. Al. 2021 | Egypt | 40 | 24 | 16 | 56.4 | 48 | Multimodal Intravenous Analgesia | 20 | Erector spinae plane block | 20 | Bilateral continuous erector spinae block provided safer and more effective analgesia, reducing opioid consumption, facilitating early extubation, and shortening ICU stay without significant adverse events. | High |
| 28 | Sharma et. Al. 2024 | India | 24 | 12 | 9 | 50.12 | 48 | Thoracic Epidural Analgesia | 12 | Erector spinae plane block | 12 | The study concluded that **the ultrasound-guided bilateral erector spinae block (ESP)** provided effective pain relief, comparable to thoracic epidural analgesia (TEA), with fewer complications. | High |
| 29 | Disikirik et. Al. 2024 | Turkey | 50 | 25 | 25 | 50 | 48 | Erector spinae plane block | 25 | Control | 25 | Preemptive ESP block reduces postoperative opioid demand and pain scores in coronary artery bypass graft surgery patients. | High |
| 30 | Sarhan et. Al. 2024 | Egypt | 219 | 139 | 80 | 42.2 | 24 | Erector spinae plane block | 109 | Fentanyl Infusion | 110 | The extubation time was halved in patients who received the single-shot bilateral ESPB compared to those who received fentanyl infusion. | High |
| 31 | Elbardan et. Al. 2024 | Egypt | 100 | 59 | 51 | 59 | 24 | Erector spinae plane block | 50 | PIFPB | 50 | Both ESPB and PIFPB are effective in enhancing recovery after sternotomy, with ESPB showing superior pain relief postoperatively and a lower requirement for opioids. | High |
| 32 | Kodali et. Al. 2022 | India | 130 | 99 | 32 | 60.19 | 24 | Erector spinae plane block | 65 | Intravenous Dexmedetomidine | 65 | The study concluded that **erector spinae fascial plane blocks** reduced postoperative pain, opioid consumption, and ICU stay duration compared to intravenous dexmedetomidine in patients undergoing off-pump coronary artery bypass grafting. | High |
| 33 | Athar et. Al. 2021 | India | 30 | 25 | 5 | 54.9 | 24 | Erector spinae plane block | 15 | Control | 15 | Ultrasound-guided Erector Spinae Plane Block (ESPB) significantly reduced 24-hour postoperative analgesic consumption by 64.5% compared to the sham block. | High |
| 34 | Hoogma et. Al. 2023 | Belgium | 64 | 55 | 9 | 64 | 24 | Erector spinae plane block | 32 | Saline | 32 | Adding an ESP block to a standard multimodal analgesia regimen did not reduce opioid consumption or pain scores following RAMIDCAB surgery. | High |
| 35 | Hoogma et. Al. 2023 | Belgium | 72 | 51 | 21 | 66 | 24 | Erector spinae plane block | 36 | Saline | 36 | Adding an ESP block to a standard multimodal analgesia regimen did not reduce opioid consumption or pain scores in minimally invasive mitral valve surgery (MIMVS). | High |
| 36 | Demir et. Al. 2024 | Turkey | 70 | 16 | 54 | 60.34 | 24 | Erector spinae plane block | 35 | Control | 35 | Using **30 mL** (vs 20 mL) per side for bilateral ESPB in CABG led to **less pain**, **vastly lower rescue tramadol**, and a **longer time to first-rescue analgesia** within 24 h. | High |
|  | Total |  | **1687** | **1073** | **637** | **55.72 ± 6.40** | **1074** |  | **856** |  | **837** |  |  |

ESP + S-PIP - Erector Spinae Plane + Superficial Parasternal Intercostal Plane, PECS + ESP - Pectoral Nerve Block + Erector Spinae Plane, PIFPB - Pecto-Interfascial Plane Block

Figure S1. Risk of bias table

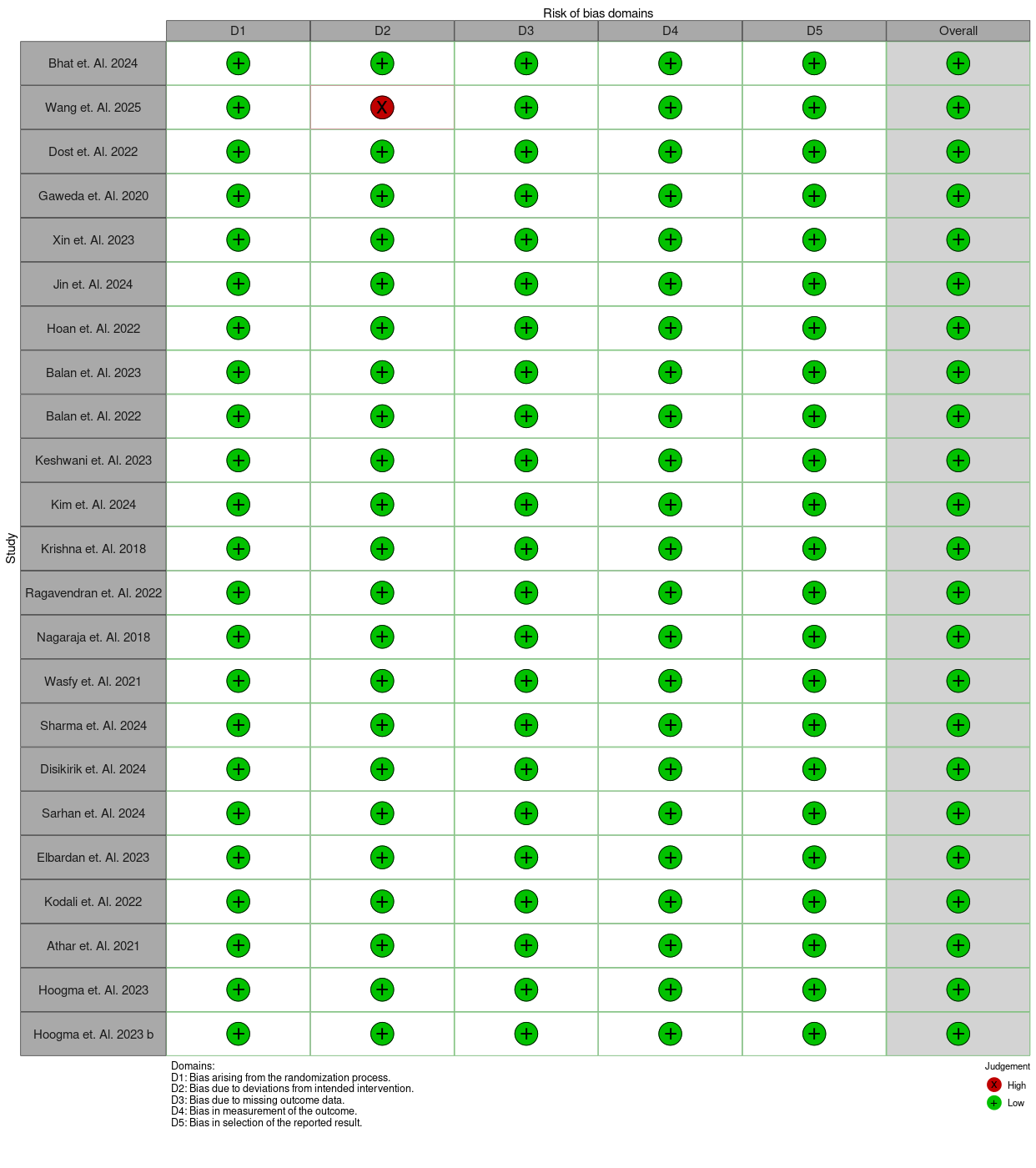

Search String

**PubMed (RCTs only)**

(("Erector Spinae Plane Block"[tiab] OR ESPB[tiab] OR ("erector spinae"[tiab] AND (block*[tiab] OR plane[tiab])) OR "Fascial Plane Block"[tiab]))
AND ("Cardiac Surgical Procedures"[MeSH] OR "cardiac surgery"[tiab] OR cardiothoracic[tiab] OR sternotomy[tiab] OR "open-heart"[tiab] OR CABG[tiab] OR "coronary artery bypass"[tiab] OR "valve surgery"[tiab])
AND (analges*[tiab] OR "postoperative pain"[MeSH] OR opioid*[tiab] OR morphine[tiab] OR safety[tiab] OR complication*[tiab])
AND (Randomized Controlled Trial[Publication Type])
Filters: Humans; Adults; English; 2025.

**Embase (Ovid; RCT limiter)**

(‘erector spinae plane block’/exp OR ESPB:ti,ab,kw OR (erector NEAR/3 spinae NEAR/3 block*):ti,ab,kw OR ‘fascial plane block’:ti,ab,kw)
AND (heart surgery/exp OR (cardiac OR cardiothoracic OR sternotomy OR ‘open-heart’ OR CABG OR ‘coronary artery bypass’ OR ‘valve surgery’):ti,ab,kw)
AND (analges* OR pain OR opioid* OR morphine OR safety OR complication*):ti,ab,kw
AND [adult]/lim AND [human]/lim
AND (‘randomized controlled trial’/de)
Limits: English; 2025.

**Scopus (TITLE-ABS-KEY; RCT terms required)**

TITLE-ABS-KEY("erector spinae plane block" OR ESPB OR (erector W/2 spinae W/2 block*) OR "fascial plane block")
AND TITLE-ABS-KEY(card* W/2 surg* OR cardiothoracic OR sternotomy OR "open-heart" OR CABG OR "coronary artery bypass" OR "valve surgery")
AND TITLE-ABS-KEY(analges* OR "postoperative pain" OR opioid* OR morphine OR safety OR complication*)
AND TITLE-ABS-KEY(random* OR "randomized controlled trial" OR placebo OR "double blind" OR "single blind")
AND (PUBYEAR < 2025)
AND (LIMIT-TO(DOCTYPE, "ar")) AND (LIMIT-TO(LANGUAGE, "English"))

**Web of Science (TS=; RCT terms required)**

TS=("erector spinae plane block" OR ESPB OR (erector NEAR/2 spinae NEAR/2 block*) OR "fascial plane block")
AND TS=(card* NEAR/2 surg* OR cardiothoracic OR sternotomy OR "open-heart" OR CABG OR "coronary artery bypass" OR "valve surgery")
AND TS=(analges* OR "postoperative pain" OR opioid* OR morphine OR safety OR complication*)
AND TS=(random* OR placebo OR "randomized controlled trial" OR "double blind" OR "single blind")
Refine: Document Type=Article; Years=2025; Language=English.

**Cochrane CENTRAL (Trials; RCT terms)**

("erector spinae plane block" OR ESPB OR (erector NEAR/3 spinae NEAR/3 block*) OR "fascial plane block"):ti,ab,kw
AND (card* NEAR/3 surg* OR sternotomy OR CABG OR "coronary artery bypass" OR "valve surgery"):ti,ab,kw
AND (random* OR placebo OR "double blind" OR "single blind"):ti,ab,kw
Limits: CENTRAL only; Adults; English;
